# Phenotype-specific associations of mosaic chromosomal alterations in systemic sclerosis

**DOI:** 10.64898/2026.03.02.26347384

**Authors:** Yuki Nishio, Yuki Ishikawa, Shunsuke Uchiyama, Xiaoxi Liu, Seiya Takada, Tomoki Kuroshima, Hajime Yoshifuji, Masanari Kodera, Mitsuteru Akahoshi, Hiroaki Niiro, Sei-ichiro Motegi, Minoru Hasegawa, Yoshihide Asano, Shingo Nakayamada, Yoshiya Tanaka, Yuriko N Koyanagi, Keitaro Matsuo, Yasushi Kawaguchi, Masataka Kuwana, Issei Imoto, Yukie Yamaguchi, Chikashi Terao

## Abstract

**Objectives:** Mosaic chromosomal alterations (mCAs) increase with age and are associated with many diseases, including autoimmune diseases. The associations between mCAs and systemic sclerosis (SSc) and its clinical subtypes have not been explored.

**Methods:** We recruited study subjects from two independent datasets (Set 1: 635 SSc, 4,401 controls; Set 2: 347 SSc, 2,170 controls) and detected mCAs (Loss, LOH, Gain, and mLOX) from their peripheral blood samples. Logistic regression analyses were conducted with covariates in each cohort, and the results were meta-analyzed. We also conducted stratified analyses by age groups, the age at disease onset, clinical phenotypes based on the skin lesions, autoantibody profiles, the presence of complications.

**Results:** We observed a trend of increased Loss in SSc, especially in old age (P=0.0063). The association of Loss was strengthened in certain subtypes of SSc, including lcSSc (OR=2.22, P=0.019) and SSc with vascular complications (digital ulcers, pulmonary hypertension, or renal crisis, OR=3.30, P=0.0054). The effect sizes of Loss increased in patients with high cell fractions (CFs). We also observed that mLOX was significantly associated with SSc, lcSSc, and ACA-SSc only for subjects with high CFs. mLOX was significantly associated with lcSSc and ACA-SSc even compared with dcSSc and ATA-SSc, respectively. These associations were consistently observed in each of the two data sets. Finally, we identified majority of the associations of Loss were mainly driven by SSc with late age at onset.

**Conclusions:** Loss and mLOX were significantly and differentially associated with SSc and its subtypes, underscoring potential phenotype-specific contributions of mCAs.

**WHAT IS ALREADY KNOWN ON THIS TOPIC:**

- Systemic sclerosis (SSc) is a heterogeneous disease, with its phenotypes and disease outcomes varying among patients.
- Age-related mosaic chromosomal alterations (mCAs) in blood and subsequent clonal haematopoiesis are associated with various adverse health outcomes.
- mCAs have also been linked to several immune-mediated diseases, such as LORA, and hence may influence immune cells and their functions.

**WHAT THIS STUDY ADDS:**

- Autosomal copy-number loss (Loss) is increased in SSc in aged subjects.
- Loss was associated with lcSSc, ACA-SSc. ILD-SSc, and VC-SSc in a dose-dependent manner of cell fraction.
- mLOX was associated with SSc and its subtypes only in patients with high cell fraction.
- Late-onset SSc and its subtypes show stronger associations with Loss with higher effect sizes compared to non-late onset SSc.

**HOW THIS STUDY MIGHT AFFECT RESEARCH, PRACTICE OR POLICY:**

- Our study facilitates further research to recapitulate the current findings in independent cohorts as well as in different ancestries.
- Incorporating profiles of Loss and mLOX in blood into conventional clinical information may enable a better stratification of SSc patients and the development of a better management strategy.
- Further experimental approaches, such as whole genome sequences and single-cell

RNA sequences, that investigate the underlying molecular mechanisms of phenotypic heterogeneity of SSc driven by Loss and mLOX are also warranted.

## Introduction

Systemic sclerosis (SSc) is a complex disease characterized by progressive fibrosis of the skin and internal organs and vasculopathy, which is thought to be caused by abnormal immune activation (1,2). SSc affects predominantly females and presents heterogeneous phenotypes based on the organ involvement (limited cutaneous, lcSSc; diffuse cutaneous, dcSSc) and serological profiles (anti-centromere antibody [ACA], anti-topoisomerase I antibody [ATA]), which are closely related to disease outcomes and hence prognosis of affected individuals (1,2). Clinical manifestations that significantly impact the prognosis include interstitial lung disease (ILD), pulmonary arterial hypertension (PAH), and other vascular manifestations (digital ulcers (DU), scleroderma renal crisis (SRC)) (1–3). For SSc patients, ILD and vascular complications can be fatal (3), and the current international guideline emphasizes early assessment of organ involvement and phenotype-based management (4). However, in clinical settings, we still often encounter treatment-resistant cases or cases with adverse effects due to intensive immunosuppressive treatments (1,2,5,6).

Genetic and environmental factors interact in the development of SSc (7). Twin studies estimated the disease heritability to be ∼4.7% (8), and the recent genome-wide association studies (GWAS) for Europeans and East Asians suggested that heritability explained by common SNPs was 0.5% and 1.3% in EUR (9) and EAS (10), respectively, indicating the relatively low heritability compared to other autoimmune diseases, such as rheumatoid arthritis (RA). Well-known environmental factors, including silica exposure (11) and chemotherapeutic agents (12) cannot fully explain the susceptibility to SSc, implicating the presence of additional acquired contributors.

Mosaic chromosomal alterations (mCAs) are acquired structural changes in autosomal and sex chromosomes and include copy loss (Loss), copy gain (Gain), copy-neutral loss of heterozygosity (CN-LOH), mosaic loss of the X chromosome (mLOX), and mosaic loss of the Y chromosome (mLOY) (13–21). It is known that mCAs increase with age, and several risk factors have also been well documented, such as cigarette smoking (13–23). Large-scale GWASs have also identified associations between genetic variants and specific mCAs (13,19–21). Beyond their role as a signature of aging, mCAs have been linked to multiple diseases, especially haematologic malignancies (13,15–18). Considering the impacts on clone sizes of B and T lymphocytes, or immune cell functions, it is expected that mCAs can also affect susceptibility to and severity of certain immune-related diseases (18).

Indeed, we have recently identified a significant correlation between mLOY and late-onset RA (LORA), RA characterized by the development at age ≥ 60 years old, a higher proportion of male patients, and clinical features distinct from young-onset RA (YORA) (24). In contrast, YORA showed a negative association with mLOY, implicating that mLOY can play distinct roles between LORA and YORA in their pathology (24). Regarding SSc, SSc typically affects individuals between 20 and 50 years of age (25). However, 10–20% of SSc cases develop in patients aged 60–65 years or even older, and the clinical features of such late-onset SSc differ from those of non-late-onset SSc (25–27). Thus, an age-at-onset-dependent association of mCA in SSc is worth investigating. Not only the age of onset, but also heterogeneity of clinical phenotypes in SSc have not been well explained by genetics as well as environmental factors thus far (9,10).

To fill the gaps, here we quantify mCA in peripheral blood obtained from subjects with or without SSc. We investigate associations of mCA with SSc and its clinical subtypes, autoantibody profiles, and major clinical manifestations relevant to prognosis. We also examine the dose-dependent effect of mCA burdens based on cell fractions (estimated proportion of peripheral-blood cells carrying given mCAs). Additionally, we investigate the age-at-onset-dependent differences of the effect sizes among SSc subjects, focusing on late-onset or non-late-onset SSc.

## Methods

### Study participants and study design

A total of 1256 cases were enrolled from multiple centers. A total of 6736 controls were enrolled from Hospital-based Epidemiological Research Program at Aichi Cancer Center (HERPACC) (28). All the study subjects were Japanese. The diagnosis of SSc was made by physicians according to the ACR/EULAR classification criteria (2013) (29). The presence of interstitial lung disease (ILD) was based on chest roentgenograms and/or CT scan images and determined in individual institutions. Patients with either pulmonary arterial hypertension (PAH), digital ulcers (DU), or renal crisis (SRC) were classified as those with vascular complications (VC). Late-onset SSc was defined as SSc with onset at age 60 years or older.

### Genotyping

Genomic DNA was extracted from peripheral blood samples and genotyped using Illumina Infinium CoreExome Array or Illumina Human CoreExome Array or a combination of Illumina Infinium Core Array and Illumina Infinium Exome Array.

### Variant- and sample-level quality control

We performed variant- and sample-level quality control (QC) before phasing for calling of mosaic events **(Figure 1A, Supplementary Figure S1)** as described in our previous work (24). Following the Mosaic Chromosomal Alterations (MoChA) WDL protocols (13,30), we excluded variants that met any of the following criteria: genotyping call rate <97%, variants falling in segmental duplications with low divergence (<2%), variants with excess heterozygosity (p<1e-6, Hardy-Weinberg equilibrium test). In sample QC, we removed genetically identical samples showing PIHAT>0.9 estimated by PLINK 1.9 (31) and outliers from East Asian clusters after we mapped the samples with the 1000 Genomes Project (32) using principal component analysis. We also removed subjects with a sample call rate < 0.97 and samples with phased B-allele frequency (BAF) autocorrelation>0.03, which indicates poor quality or contamination of DNA. The final cleaned mosaic call set included Set 1, consisting of 635 SSc cases and 4,401 non-SSc controls, and Set 2, consisting of 347 SSc cases and 2,170 non-SSc controls **(Figure 1B)**.

**Figure 1.**
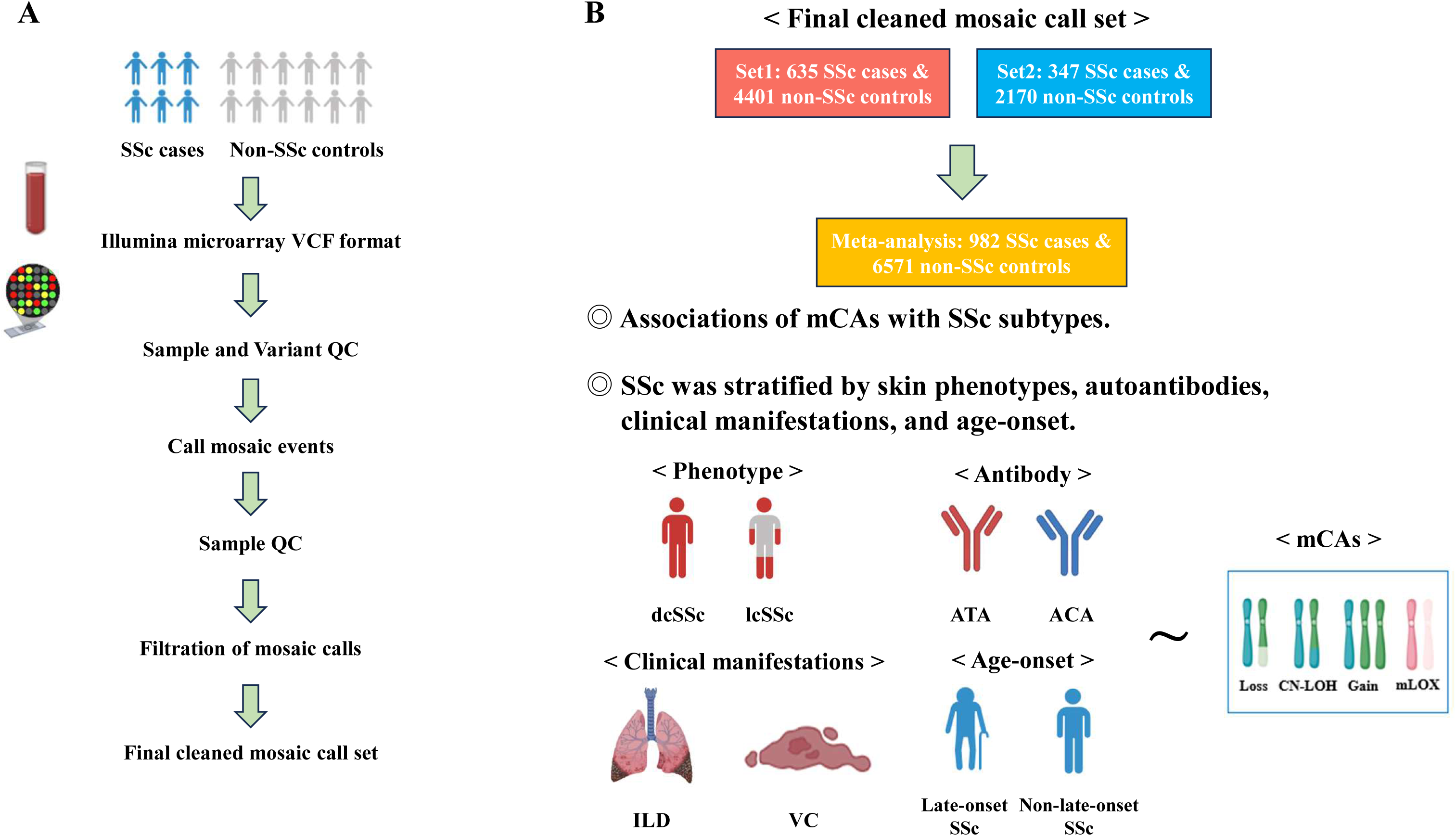
Overview of the present study. (A) A workflow of the mosaic call. (B) The design of the present study. SSc, systemic sclerosis; mCA, mosaic chromosomal alteration; CN-LOH, copy-neutral loss of heterozygosity; mLOX, mosaic loss of chromosome X; dcSSc, diffuse cutaneous SSc; lcSSc, limited cutaneous SSc; ATA, anti–topoisomerase I antibody; ACA, anti-centromere antibody; ILD, interstitial lung disease; VC, vascular complications; QC, quality control; VCF, Variant Call Format.

### Mosaic event detection

Mosaic call detection was performed as described in our previous work (24) and summarized in Figure 1B. After sample- and variant-level QCs, the long-range haplotype phasing was performed using SHAPEIT5 (33). mCAs were detected using MoChA WDL pipelines (2024 09-27 version) (13,30). After running MoChA, we excluded subjects that met one of the following criteria: subjects aged less than 16 years old at the time of sampling, those with sex discordance between genotypes and phenotypes, or subjects with missing data on their sex and/or age. Mosaic calls were filtered by excluding CNP-class events and retaining only calls supported by BAF- and coverage/length-based criteria (thresholds on bdev, n50_hets, lod_BAF metrics, rel_cov, and event length). Regarding autosomal events, we excluded constitutional mosaic events (lod_baf_phase <10), and germline duplications (length < 500 kbp and relative coverage > 2.5). To reduce potential sex-chromosome aneuploidy, mLOX events were restricted to events with relative coverage < 2.5. Autosomal mCA events were detected by copy number state (Loss, CN-LOH, or Gain) and by p versus q arm for loss and CN-LOH events. mCAs for which we could not determine the copy number state were categorised as undetermined and included in mCAs. mLOX was detected only in females. For each Loss and mLOX that passed QCs, cell fraction (CF) was calculated as 4×bdev/(1+2×bdev), where bdev represents the BAF deviation estimated from 0.5.

### Statistical analysis

We performed logistic regression analyses to evaluate the association between SSc and mCAs as follows,

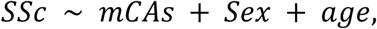

where mCAs are any autosomal mCA, Loss, LOH, Gain, or mLOX. Only female samples were analysed for mLOX without sex as a covariate. In SSc, women are the majority, so we did not perform the mLOY analysis.

We tested the association of Loss or mLOX with SSc subtypes as follows,

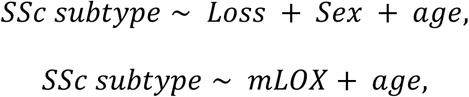

where subjects were stratified by skin phenotypes (lcSSc, dcSSc), autoantibody profiles (ACA, ATA), the presence of ILD or VC (only Loss), or ages (≥60, <60, only Loss). Intra-case analyses of associations between mLOX and skin phenotypes or autoantibody profiles were also conducted using this model.

For conditional analyses, the associations of SSc subtypes and Loss were conditioned on the skin phenotypes or autoantibody profiles,

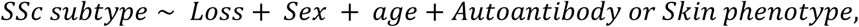

where SSc subtypes were ILD or VC.

To test the dose-dependent effect of CF, we compared subjects who carried Loss or mLOX with CF> 5% and subjects without any mCAs,

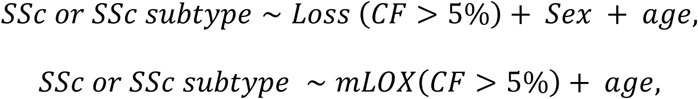

where subjects were stratified by skin phenotypes (lcSSc), autoantibody profiles (ACA), or the presence of ILD or VC (only Loss).

For association analyses of the age of disease onset, we tested the association of Loss or mLOX with late-onset or non-late-onset SSc or its subtypes (lcSSc, ACA(+), ILD(+) or VC(+)) in the following model.

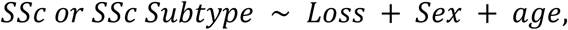

All the analyses were performed separately in the two datasets (Sets 1 and 2), and then the results were meta-analyzed (**Figure 1B**). Meta-analyses were performed using an inverse variance fixed-effects model. In the case of complete separation, associations were estimated using Firth regression, and inference was performed using profile likelihood.

Statistical analyses were conducted using R (version 4.5.0) and Python (version 3.11.8). The estimated risk is expressed as an odds ratio (OR) with its 95% confidence interval (CI). The statistical significance of the associations were defined based on Bonferroni’s correction.

### Ethical and public involvement statements

All patients provided informed consent. This study was approved by the local ethics committee of each institute as previously described (10). Patients were not involved in any part of this study, including the study design, formal analyses, result interpretation, writing, or editing of the manuscript.

## Results

### Demographic and clinical features of study participants

The demographic features of the participants in Sets 1 and 2 are listed in **Table 1**. As expected, SSc patients included more females than controls in both datasets.

**Table 1.** Demographic features of the subjects in each dataset.

| Datasets | Set 1 |  | Set 2 |  |
| --- | --- | --- | --- | --- |
|  | SSc | Controls | SSc | Controls |
| **Number of subjects,** | **635** | **4401** | **347** | **2170** |
| **age in median (IQR)** | **56 (42–65)** | **57 (47–64)** | **65 (54–72)** | **57 (47–64)** |
| **Sex, n (%)** |  |  |  |  |
| **Male** | **69 (10.9%)** | **2552 (58.0%)** | **51 (14.7%)** | **1259 (58.0%)** |
| **Female** | **566 (89.1%)** | **1849 (42.0%)** | **296 (85.3%)** | **911 (42.0%)** |
| **Skin phenotype, n (%)** |  |  |  |  |
| **lcSSc** | **381 (60.0%)** | **—** | **231 (66.6%)** | **—** |
| **dcSSc** | **254 (40.0%)** | **—** | **115 (33.1%)** | **—** |
| **Autoantibody, n (%)** |  |  |  |  |
| **ACA** | **225 (35.4%)** | **—** | **169 (48.7%)** | **—** |
| **ATA** | **161 (25.4%)** | **—** | **76 (21.9%)** | **—** |
| **Clinical manifestations, n (%)** |  |  |  |  |
| **ILD** | **289 (45.5%)** | **—** | **143 (41.2%)** | **—** |
| **VC** | **176 (27.7%)** | **—** | **88 (25.4%)** | **—** |
| **Age at onset, n (%)** |  |  |  |  |
| **Late-onset** | **56 (8.8%)** | **—** | **121 (34.9%)** | **—** |
| **Non-late-onset** | **132 (20.8%)** | **—** | **222 (64.0%)** | **—** |

SSc, systemic sclerosis; dcSSc, diffuse cutaneous systemic sclerosis; lcSSc, limited cutaneous systemic sclerosis; ATA, anti-topoisomerase I antibody; ACA, anti-centromere antibody; ILD, interstitial lung disease; VC, vascular complications; IQR, interquartile range; Late-onset of the disease is defined as the disease onset at the age $\geq 60$ years old.

Consistent with previous reports (13–21), the prevalence of mCAs increases with age in both SSc and control cohorts, confirming the validity of the current results (**Supplementary Figure S2, Supplementary Table S1)**.

### An age-dependent positive association of Loss with SSc

To investigate if the presence of specific types of mCAs is associated with SSc, we applied the logistic regression models with covariates (**Methods**). We identified a trend of increased mCAs in SSc and this was driven by consistent positive associations between Loss and SSc across the datasets (OR = 1.72; 95% CI, 0.94-3.14; P = 0.079, meta-analysis) (**Figure 2A, Supplementary Table S2**). Thus, we primarily focused on Loss in the following analyses. Though it is well-known that mCAs increased with age even in healthy individuals (13–21) (**Supplementary Figure S2, Supplementary Table S1**), we observed that the effect sizes of Loss on SSc were significantly higher in patients aged 60 years or more (OR = 3.02; 95% CI, 1.37-6.66; P = 0.0063), which was consistent across the datasets (**Figure 2B, Supplementary Table S3**).

**Figure 2.**
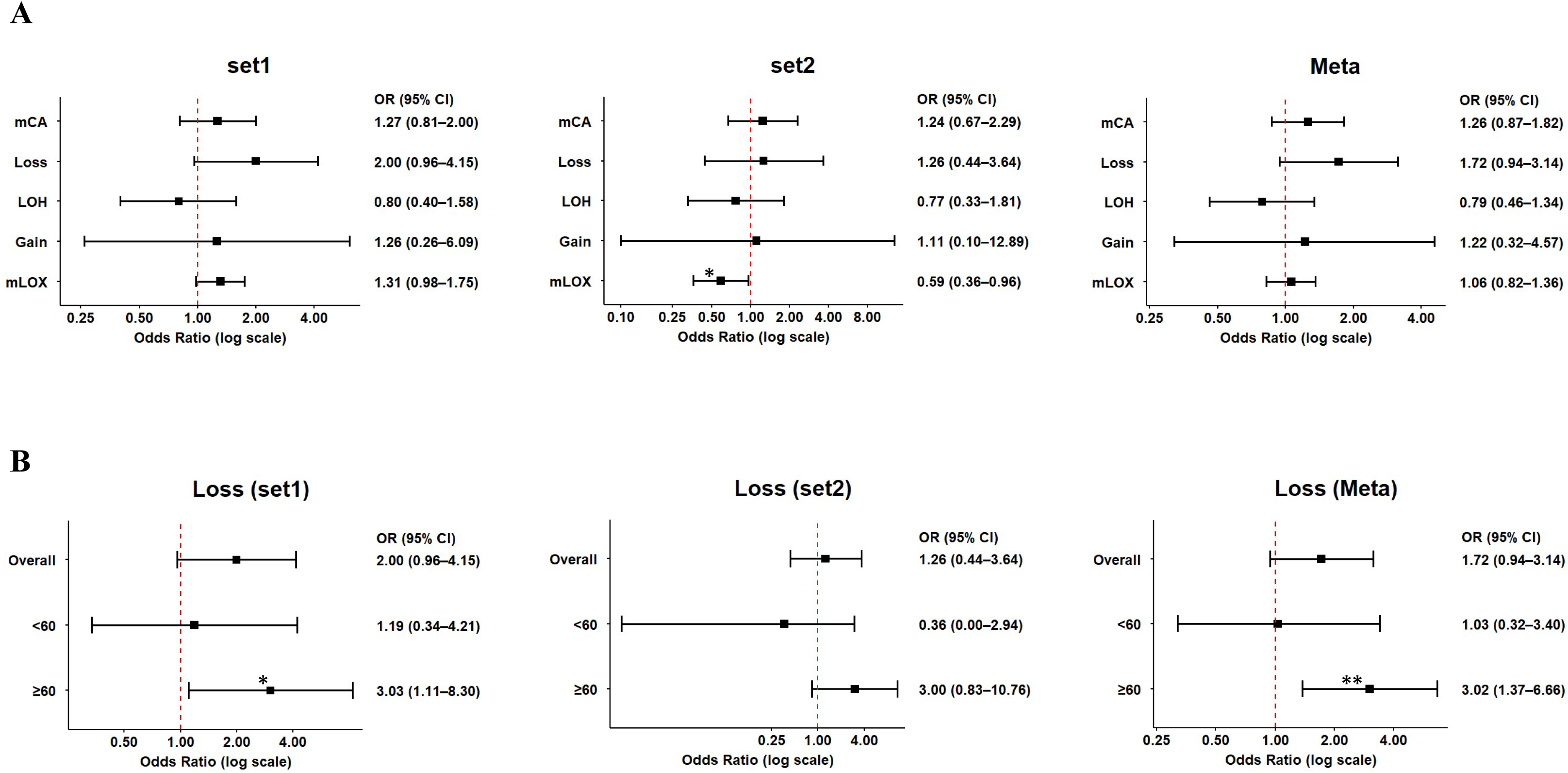
Association between SSc and mCAs. (A) Forest plots showing associations between SSc and mCAs or its subtypes (Loss, LOH, Gain, and mLOX) in two independent datasets (Set 1, Set 2) and those of meta-analysis are presented. (B) Forest plots showing associations between SSc and Loss across different age groups (age < 60 and age ≥ 60 years) in two independent datasets (Set 1, Set 2) and those of meta-analysis are presented. Odds ratios (ORs) are presented in a log scale, and error bars indicate the 95% confidence intervals (CIs). Statistical significance is denoted based on nominal two-sided p-values of P<0.05 (*) and P<0.01 (**). mCA, mosaic chromosomal alteration; LOH, Loss of heterozygosity; mLOX, mosaic loss of chromosome X.

These findings suggest that Loss is significantly associated with SSc especially for patients with advanced age.

### Phenotype-specific associations of Loss

We investigated if the heterogeneity of SSc phenotypes of SSc, such as skin manifestations (lcSSc or dcSSc) or autoantibody profiles (ACA or ATA), is associated with a specific type of mCAs. We identified a significant positive association of Loss with lcSSc (OR = 2.22; 95% CI, 1.14-4.33; P = 0.019; meta-analysis), which is consistent across the datasets (**Figure 3A, Supplementary Table S4**). We also observed consistent positive associations of Loss with ACA-SSc (OR = 2.27; 95% CI, 1.01-5.14; P = 0.049; meta-analysis) (**Figure 3B, Supplementary Table S5**). On the other hand, while dcSSc has no association with Loss, ATA-SSc shows positive association with Loss (OR = 2.97; 95% CI, 1.14-7.70; P = 0.026; meta-analysis) (**Figure 3A and B, Supplementary Table S4 and S5**).

**Figure 3.**
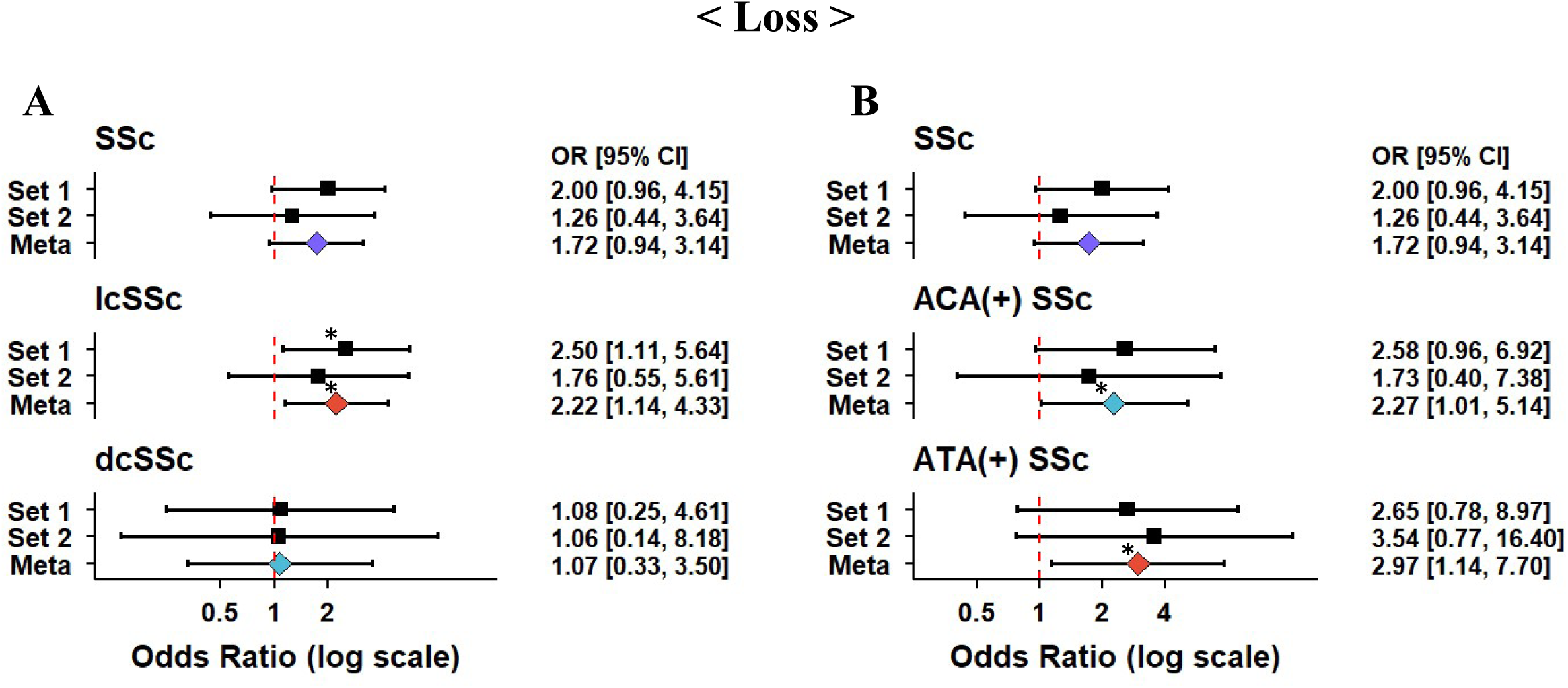
Associations of Loss with major clinical subtypes of SSc. (A-B) Associations between skin subtypes (lcSSc and dcSSc) and Loss (A) or between autoantibody profiles (ACA and ATA) and Loss (B) are presented. The associations between SSc and Loss are presented for comparison. Presented are the results of a case-control study. Odds ratios (ORs) are presented in a log scale, and error bars indicate the 95% confidence intervals (CIs). Statistical significance is denoted based on nominal two-sided p-values of P<0.05 (*). SSc, systemic sclerosis; lcSSc, limited cutaneous SSc; dcSSc, diffuse cutaneous SSc; ACA, anti-centromere antibody; ATA, anti– topoisomerase I antibody.

Then, we sought potential associations of clinical phenotypes other than skin manifestations and autoantibody profiles, namely ILD and VC, both of which are intractable to treatment and can even be fatal (3). We identified that Loss had a significant positive association with ILD (OR = 2.44; 95% CI, 1.16-5.14; P = 0.019; meta-analysis) and VC (OR = 3.30; 95% CI, 1.42-7.66; P = 0.0054; meta-analysis) (**Figure 4A and B, Supplementary Table S6**). Of note, the effect sizes tended to be larger than that of SSc and Loss. Furthermore, the associations between Loss and ILD or VC remained significant even after conditioning on the presence of ACA or ATA (**Figure 4C and D, Supplementary Table S7**). Conditioning on the skin phenotypes (lcSSc or dcSSc), the associations were attenuated; but a trend of positive association still remained (**Supplementary Figure S3, Supplementary Table S7**).

**Figure 4.**
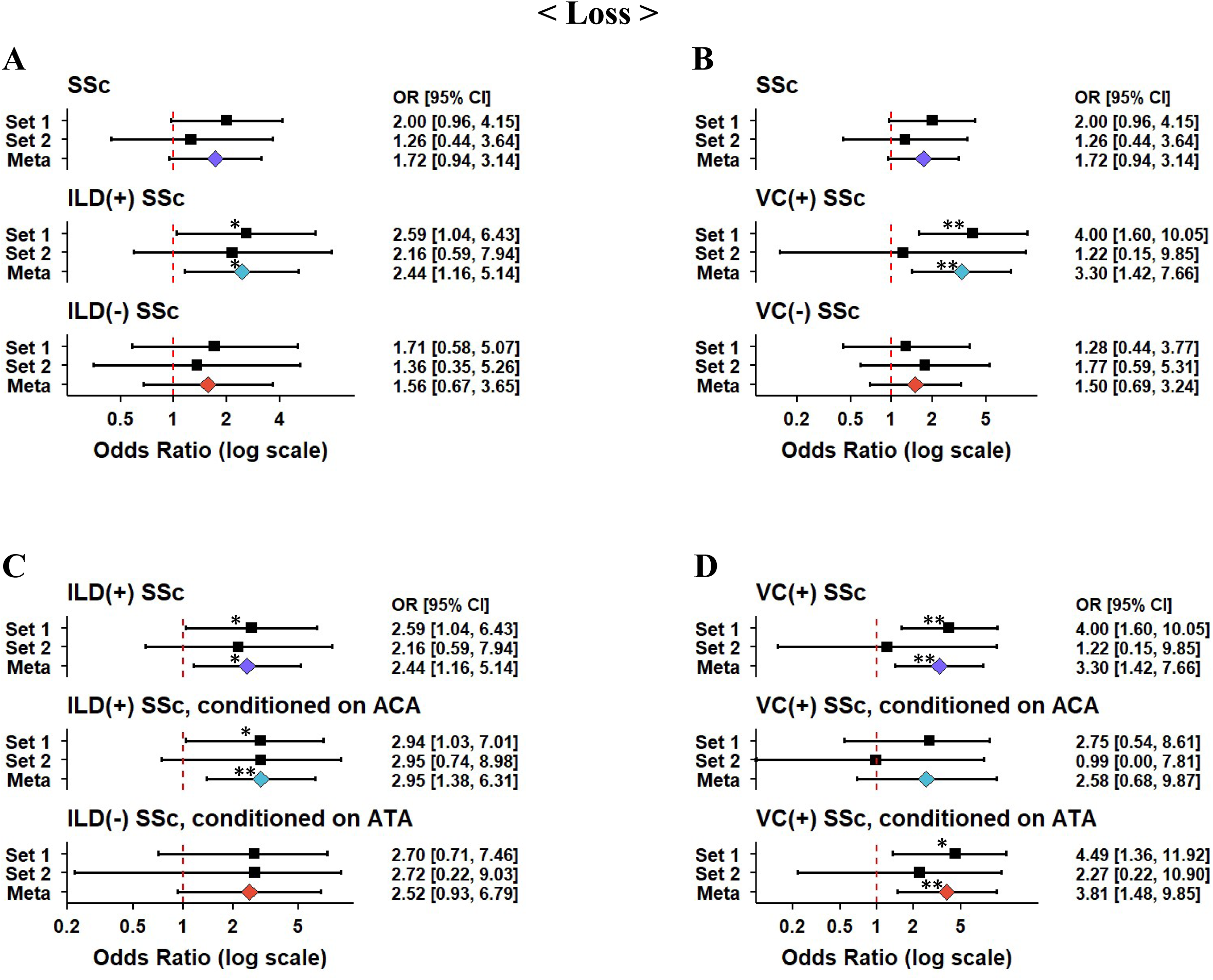
Associations between Loss and SSc with interstitial lung disease or vascular complications. (A, B) Associations between SSc with or without interstitial lung disease (ILD) and Loss (A) and SSc with or without vascular complications (VC) and Loss (B) are presented. The associations between SSc and Loss are presented for comparison. (C, D) Associations between ILD-SSc and Loss (C) or VC-SSc and Loss conditioning on autoantibody profiles (ACA, ATA) are presented. The associations between ILD-SSc or VC-SSc and Loss are presented for comparison. The results of a case-control study are shown. Odds ratios (ORs) are presented in a log scale, and error bars indicate the 95% confidence intervals (CIs). Statistical significance is denoted based on two-sided nominal p-values of P<0.05 (*) and P<0.01 (**). SSc, systemic sclerosis; ACA, anti-centromere antibody; ATA, anti–topoisomerase I antibody.

Together, these results suggest the phenotype-specific association of Loss.

### Dose-dependent associations of Loss with SSc subsets

Considering the survival advantage, and clonal expansion of the haematopoietic stem and progenitor cells (HSPCs) with given mCAs (13,34), the higher burden of mCAs can confer larger effects on phenotypes.

To test such a dose-dependent effect, we extracted the subjects with Loss with CF > 5% and compared the risk of each phenotype in a case-control setting. As we expected, we observed increased effect sizes in associations of Loss with SSc, lcSSc, ACA-SSc, ILD-SSc, and VC-SSc in subjects with high CF (**Figure 5, Supplementary Table S8**).

**Figure 5.**
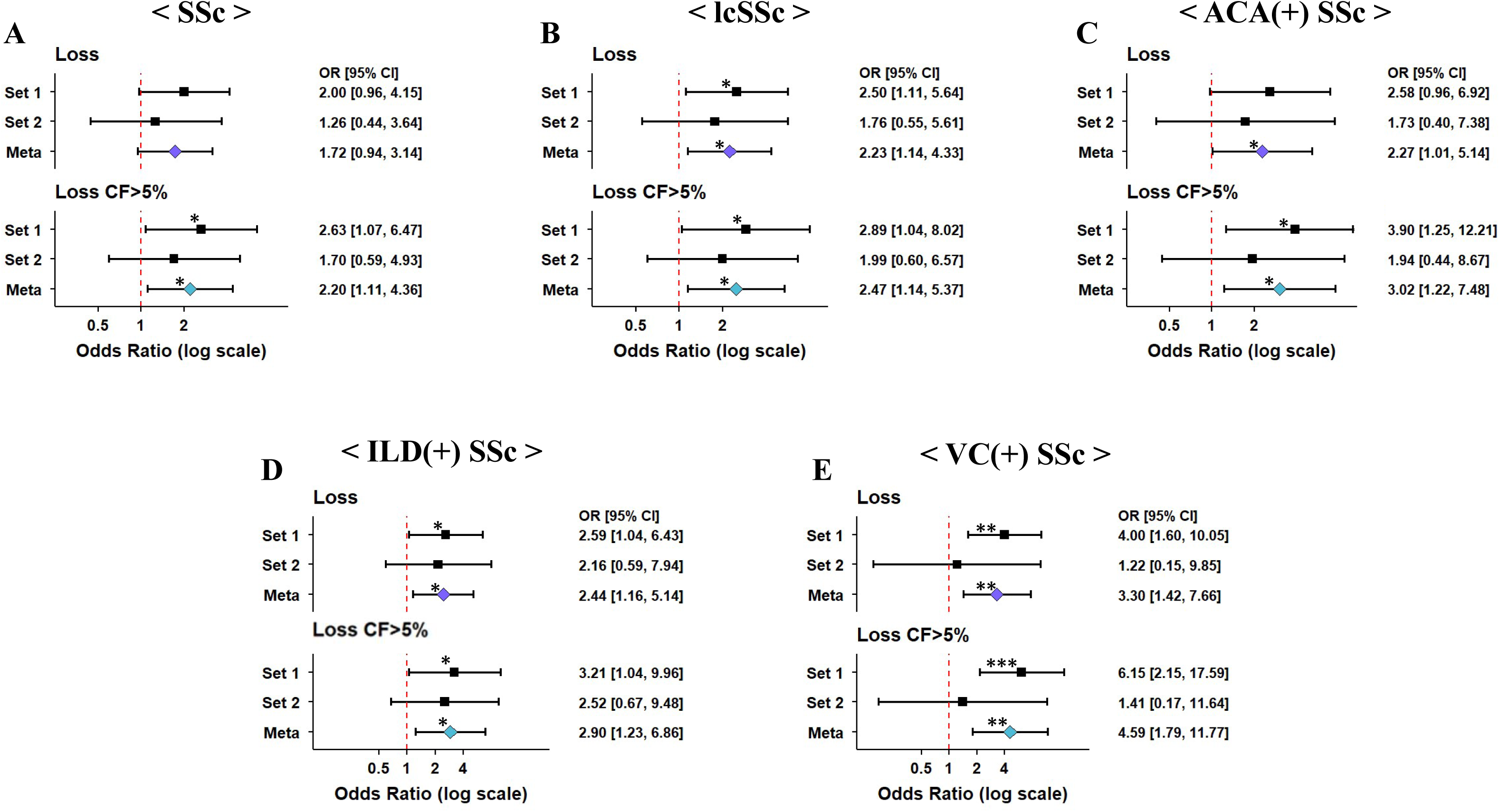
Dose-dependent effect of the cell fraction of Loss on SSc and its subtypes. (A-E) Associations between Loss and SSc (A), lcSSc (B), ACA-SSc (C), ILD-SSc (D), or VC-SSc (E) with (bottom) or without (top) cell fraction (CF) threshold of 5% (Loss CF > 5%) are presented. The results of a case-control study are shown. Odds ratios (ORs) are presented on a log scale, and error bars indicate the 95% confidence intervals (CIs). Statistical significance is denoted based on two-sided nominal p-values of P<0.05 (*), P<0.01 (**), and P<0.001 (***). SSc, systemic sclerosis; lcSSc, limited cutaneous SSc; ACA, anti-centromere antibody; ILD, interstitial lung disease; VC, vascular complications.

### Associations of mLOX were mainly observed in subjects with high CF

The increased effect sizes in subjects with high CF for Loss motivated us to analyze other types of mCAs in detail for associations only observed in subjects with high CF.

As a result, we identified consistent positive associations between mLOX with high CF and SSc (OR = 2.17; 95% CI, 1.00-4.70; P = 0.050), lcSSc (OR = 2.84; 95% CI, 1.27-6.34; P = 0.011), and ACA-SSc (OR = 3.72; 95% CI, 1.60-8.65; P = 0.0023), that were not apparent without considering CF (**Figure 6A-C, Supplementary Table S9**).

**Figure 6.**
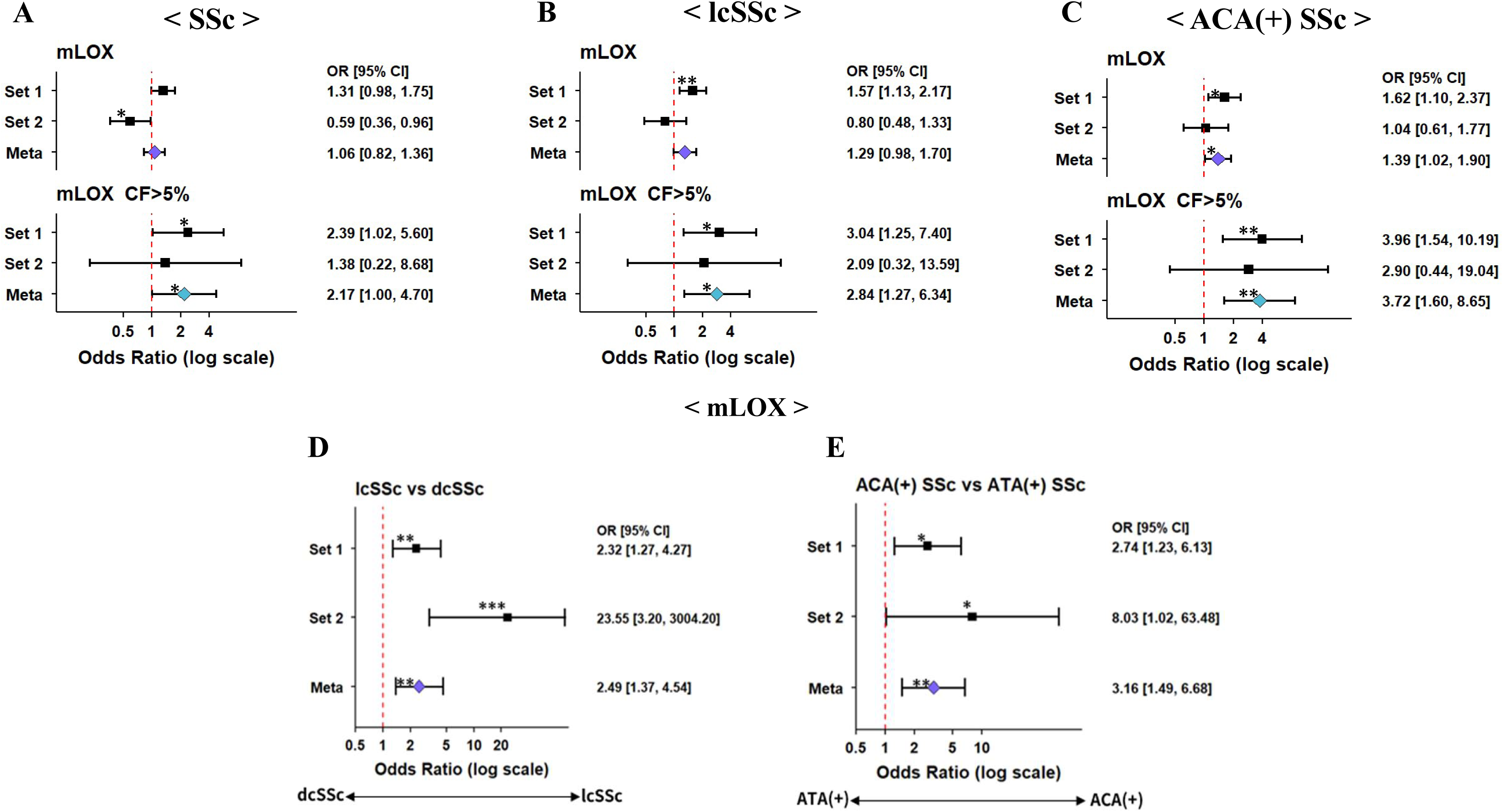
Intra-case association of mLOX with major clinical subtypes of SSc and dose-dependent effect of the cell fraction of mLOX on SSc and its subtypes. (A-C) Associations of mLOX with SSc (A), lcSSc (B), and ACA-SSc (C), with (bottom) or without (top) cell fraction (CF) threshold of 5% (mLOX CF > 5%) are presented. The results of a case-control study are shown. Odds ratios (ORs) are presented in a log scale, and error bars indicate the 95% confidence intervals (CIs). Statistical significance is denoted based on two-sided nominal p-values of P<0.05 (*), P<0.01 (**), and P<0.001 (***). SSc, systemic sclerosis; lcSSc, limited cutaneous SSc; dcSSc, diffuse cutaneous SSc; ACA, anti-centromere antibody; ATA, anti–topoisomerase I antibody; mLOX, mosaic loss of chromosome X. (D-E) Intra-case associations of mLOX comparing skin subtypes (lcSSc and dcSSc) (D) and autoantibody profiles (ACA and ATA) (E) are presented.

We also found that mLOX showed significant associations in intra-case analyses of SSc subsets, namely, positive associations of lcSSc in comparison with dcSSc (OR = 2.49; 95% CI, 1.37-4.54; P = 0.0027) and positive associations of ACA-SSc in comparison with ATA-SSc (OR = 3.16; 95% CI, 1.49-6.68; P = 0.0026) (**Figure 6D and E, Supplementary Table S10**). These effect sizes in intra-case analyses were further increased in the analyses restricting to subjects with high CF although the associations were no longer significant due to the limited sample sizes (**Supplementary Figure S4, Supplementary Table S11**). Interestingly, we did not observe significant associations of Loss in the intra-case analyses (**Supplementary Figure S5, Supplementary Table S12**).

These results suggest the dose-dependent involvement of mLOX with SSc and its subtypes.

### The stronger association of Loss with late-onset SSc and its subtypes

Based on our previous findings of LORA-specific associations, we hypothesized that there might be differential associations between subjects with different ages of disease onset. Referring to the previous studies (24,25,27), we stratified the patients with SSc or its clinical subtypes according to the age of disease onset (≥60 and <60 years) (**Methods**).

We observed stronger associations of Loss and late-onset subsets of SSc (OR = 3.94; 95% CI, 1.43-10.86; P = 0.0080), lcSSc (OR = 4.77; 95% CI, 1.48-15.35; P = 0.0088), ACA-SSc (OR = 8.13; 95% CI, 1.95-33.96; P = 0.0040), ILD-SSc (OR = 3.56; 95% CI, 0.80-15.86; P = 0.096), and VC-SSc (OR = 6.53; 95% CI, 1.35-31.60; P = 0.020) with higher effect sizes than those of overall and non-late-onset subsets. (**Figure 7, Supplementary Table S13 and S14**). On the other hand, we did not observe consistent patterns in mLOX (**Supplementary Table S15 and S16**).

**Figure 7.**
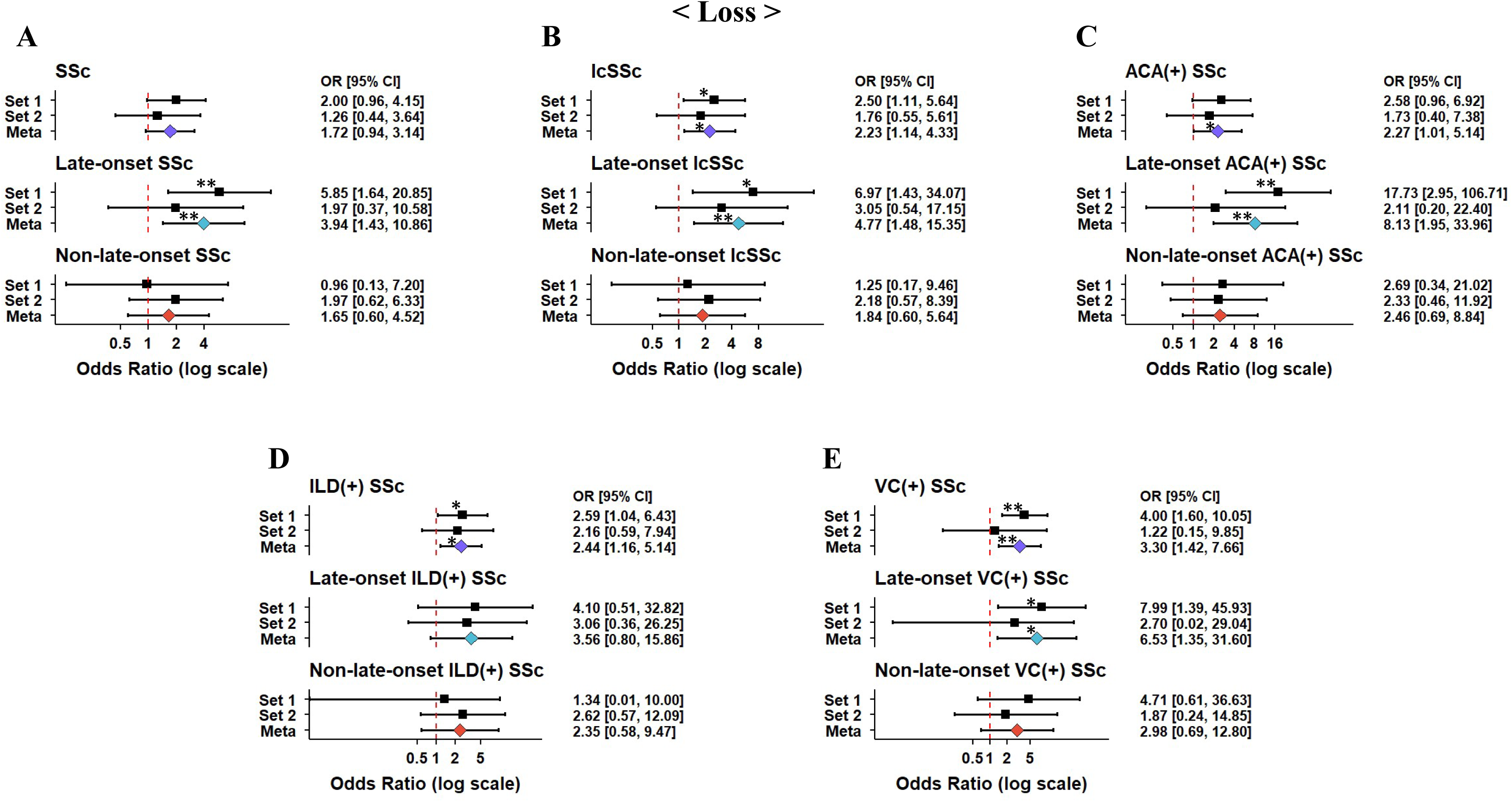
Stronger associations of Loss with late-onset SSc and its subtypes. (A-E) Associations between Loss and SSc (A), lcSSc (B), ACA-SSc (C), ILD-SSc (D), and VC-SSc (E) for the whole subset (top), the subset with late-onset (middle), and those with non-late-onset (bottom) are presented. The results of a case-control study are shown. Odds ratios (ORs) are presented on a log scale, and error bars indicate the 95% confidence intervals (CIs). Statistical significance is denoted based on two-sided nominal p-values of P<0.05 (*), P<0.01 (**). SSc, systemic sclerosis; lcSSc, limited cutaneous SSc; ACA, anti-centromere antibody; ILD, interstitial lung disease; VC, vascular complications.

These results support the notion that phenotypes of late-onset SSc are different from those of non-late-onset SSc, and Loss can partially explain such phenotypic differences.

### The Loss events in SSc tended to be skewed in chr5q

Since we observed associations of Loss with SSc and its subtypes, we analyzed whether Loss events in SSc recurrently involved a specific genomic region. We observed Loss in the long arm of chromosome 5 (5q) were the most frequent in SSc (20.0%). The 5q Loss were the second most frequent in the controls (9.18%). While this study with low statistical power did not reveal a significant result, the 5q region may be associated with SSc.

## Discussion

In the present study, we identified associations of Loss and mLOX with SSc and its subtypes. The consistent association patterns across data sets support the validity of the results. A part of associations were pronounced by stratifications based on biological age, clinical phenotypes and age at disease onset, highlighting the potential roles of mCAs in the pathologic basis of SSc and its heterogeneous phenotypes.

Although there is no established evidence how mCAs impact on immune cells so far, the present study together with our previous finding of mLOY in LORA suggest that mCAs might contribute to the expansion of aberrant immune cell clones and subsequent dysregulation of immune responses (24). Of note, previous works have suggested that somatic genome alterations play some roles in SSc biology. A study with target sequence of genes frequently mutated in myeloid malignancies in peripheral blood mononuclear cells from SSc and healthy donors identified enrichment of clonal haematopoiesis of intermediate potential (CHIP) in younger (age < 50 years) SSc patients (35). Another study with whole-genome sequence of cultured lung fibroblasts from SSc and healthy subjects demonstrated increased somatic mutational burden and chromosomal instability in SSc lung fibroblasts (36). Hence, the present study expands the landscape of somatic genome architectures in SSc by suggesting that Loss and mLOX contribute to phenotypic heterogeneity.

The association between Loss and VC suggests that such chromosomal alterations may lead to endothelial dysfunction and injury with impaired vascular repair process, leading to vascular lesions observed in SSc, including DU, PAH, and SRC. We observed the dose-dependent associations of Loss as well as associations of mLOX only in subjects with high CF, suggesting that the expansion of Loss-bearing clones (in either autosome or X chromosome) is likely to exert measurable effects (e.g., vascular impairment) leading to specific phenotype manifestations (e.g., vascular complications). Importantly, the effect sizes of Loss were pronounced in late-onset SSc, especially in lcSSc and ACA phenotypes, well concordant with the previous finding that late-onset SSc exhibits a higher prevalence of lcSSc and ACA phenotypes (25–27). These data may suggest that age-related chromosomal instability and clonal haematopoiesis of Loss underlie the phenotypic shift toward the limited form (rather than the generalized form) of skin phenotype or the vascular form (rather than the immune-fibrotic form) in late-onset SSc. Together, these data suggest that mCAs, particularly Loss and mLOX, might serve as biomarkers, partly in a dose-dependent manner, and hence contribute to the development of phenotype-based clinical management of SSc.

There are several limitations in this study, which need to be addressed in future studies. First, its cross-sectional design hinders testing causal relationships between mCAs and disease onset or progression. Second, the current study is still underpowered to detect further subtype-specific associations. Third, the effects of untested confounders, such as treatment history, environmental exposures, or unmeasured clinical variables, cannot be fully excluded.

Nevertheless, our data provide novel biological insights that illuminate unrecognized immune-haematopoietic pathways underlying clinical heterogeneity in SSc. Further expansion of data, including samples from other ancestries and sequencing data (whole-genome sequencing and single-cell RNA sequencing), would help us to consolidate the current findings and elucidate the cellular and molecular contributions of mCAs to SSc pathophysiology.

### Declaration of generative AI and AI-assisted technologies in the manuscript preparation process

During the preparation of this work, the authors used ChatGPT (OpenAI) to improve the clarity and readability of the English text. After using this tool, the authors reviewed and edited the manuscript as needed and take full responsibility for the content of the published article.

## Data Availability

All data produced in the present study are available upon reasonable request to the authors

## Acknowledgements

We thank Systemic Sclerosis Working Group of Japan Ministry of Health, Labour and Welfare for collecting and managing the samples and clinical information.

## Contributors

YN and CT conceived the project. YY and CT supervised the project. YN and CT analysed the data. YI, SU, ST, and TK shared the statistical codes. CT, HY, MK, MA, HN, SM, MH, YA, SN, YT, YK, and MK generated the SSc data. YNK, KM, and II generated the control data. CT, YI, SU, and XL participated in technical discussions of mCAs. YN, YI, and CT wrote the manuscript. All authors have critically reviewed and approved the final version of the manuscript. CT is guarantor.

## Patient consent for publication

Not applicable.

## Ethics approval

All patients provided informed consent, and this study was approved by the local ethics committee of each institute.

