## Supplementary Appendix for "Phenotype-specific associations of mosaic chromosomal alterations in systemic sclerosis"

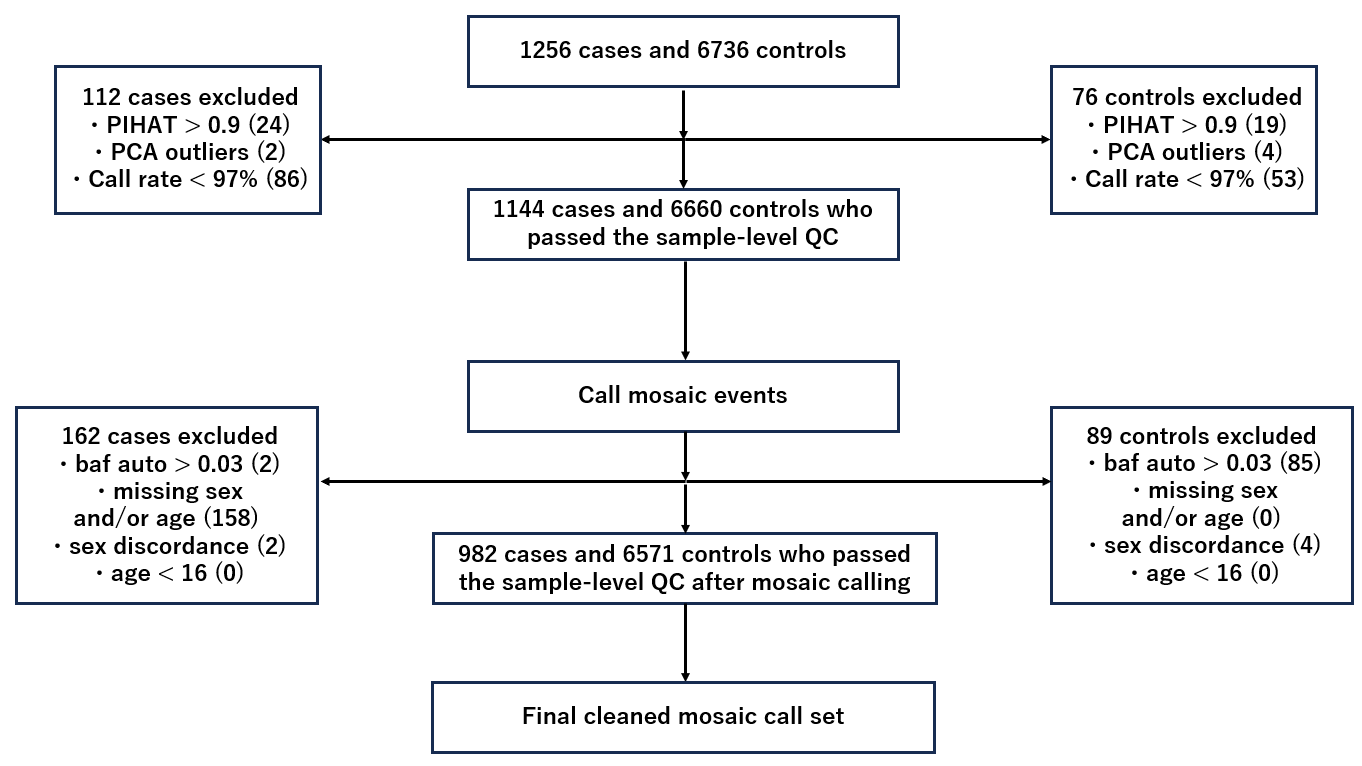

**Supplementary Figure S1. Flow chart of sample-level QC in the current study.**

The number of the samples excluded in the sample-level QC is shown in parentheses.

PCA, principal component analysis.

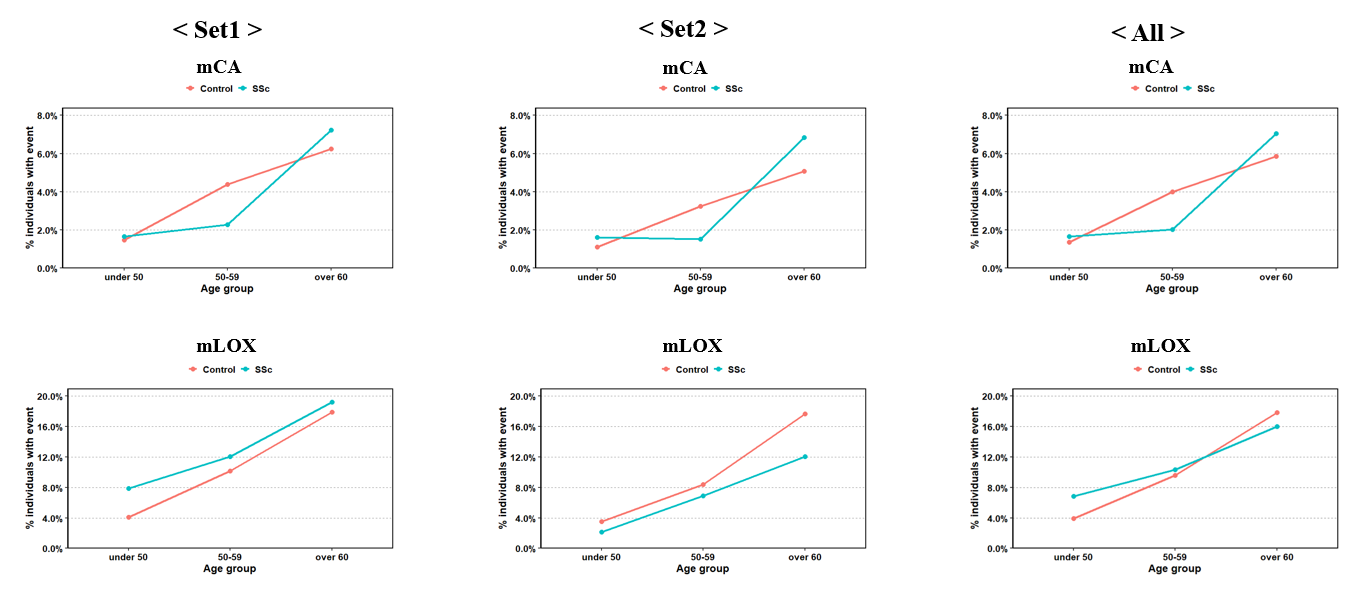

**Supplementary Figure S2. Prevalence of mosaic events in SSc and controls by different age groups.**

Percentage of individuals with any autosomal mCA and mLOX by age group (<50, 50–59, ≥60) in SSc and controls in each dataset (Set1, Set2, and All) are presented.

SSc, systemic sclerosis; mCA, mosaic chromosomal alteration; mLOX, mosaic loss of chromosome X.

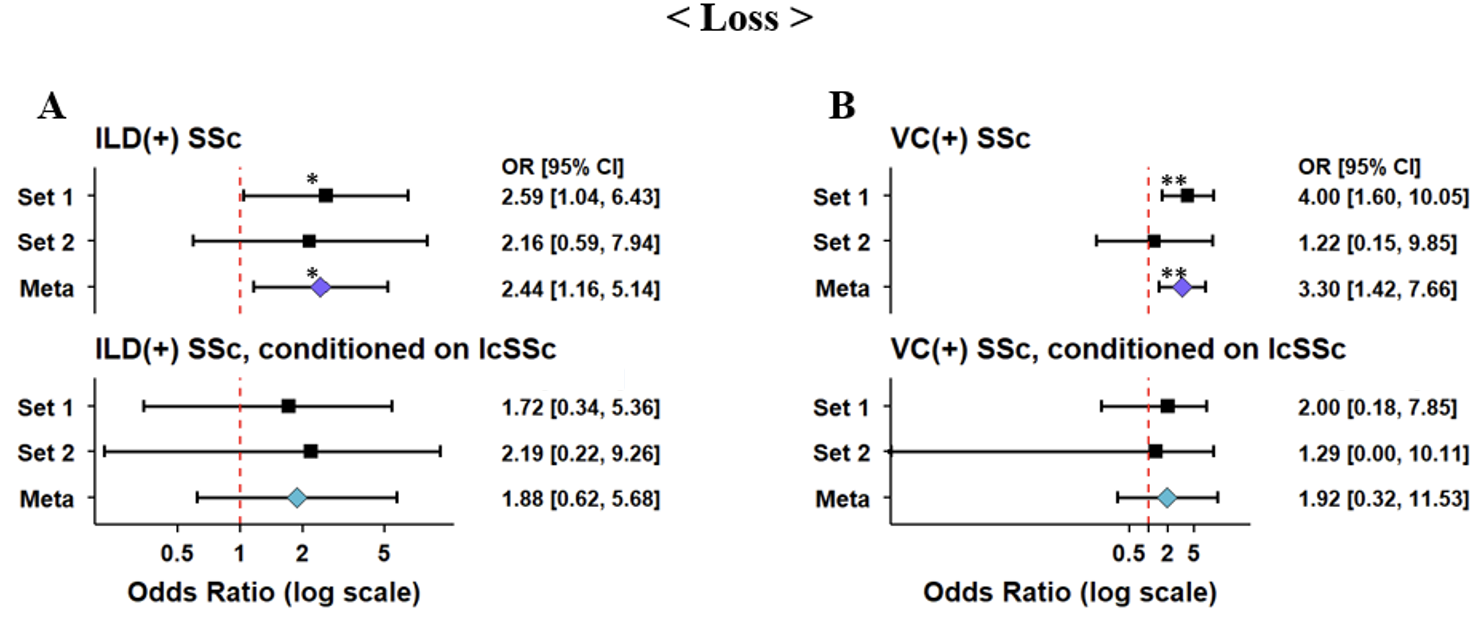

**Supplementary Figure S3. Associations between Loss and SSc with interstitial lung disease or vascular complications, conditioned on lcSSc.**

(A, B) Associations between SSc with interstitial lung disease (ILD-SSc) and Loss (A) or SSc with vascular complications (VC-SSc) and Loss (B) conditioned on skin subtypes (lcSSc) are presented, referring to associations between ILD-SSc or VC-SSc and Loss. Case-control study results of each dataset and the meta-analysis results are shown. Odds ratios (ORs) are presented on a log scale, and error bars indicate the 95% confidence intervals (CIs). Statistical significance is denoted based on two-sided nominal p-values of P<0.05 (*), and P<0.01 (**). SSc, systemic sclerosis; lcSSc, limited cutaneous SSc; ILD, interstitial lung disease; VC, vascular complications.

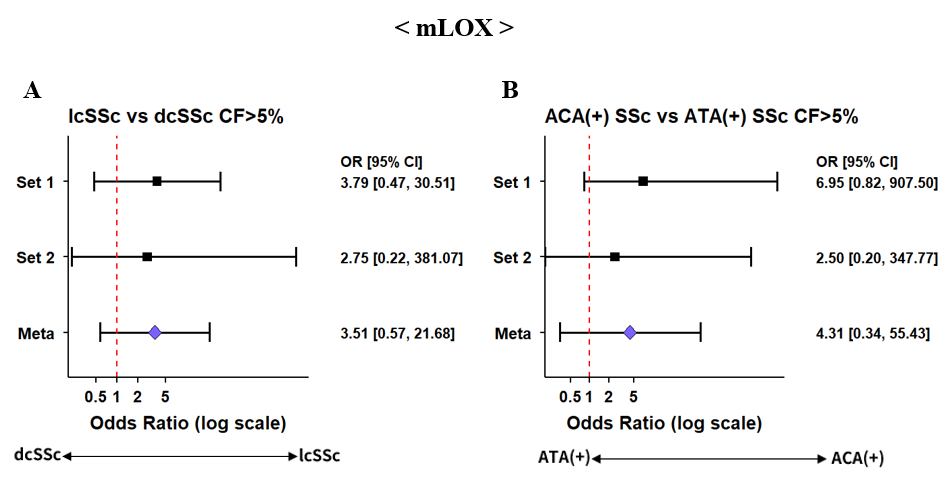

**Supplementary Figure S4. Intra-case associations between mLOX (CF>5%) and major clinical subtypes of SSc.**

Associations between skin subtypes (lcSSc and dcSSc) and mLOX (CF>5%) (A) and between autoantibody profiles (ACA and ATA) and mLOX (CF>5%) (B) are presented. Intra-case results of each dataset and the meta-analysis results are shown. Odds ratios (ORs) are presented on a log scale, and error bars indicate the 95% confidence intervals (CIs). SSc, systemic sclerosis; lcSSc, limited cutaneous SSc; dcSSc, diffuse cutaneous SSc; ACA, anti-centromere antibody; ATA, anti–topoisomerase I antibody; mLOX, mosaic loss of chromosome X.

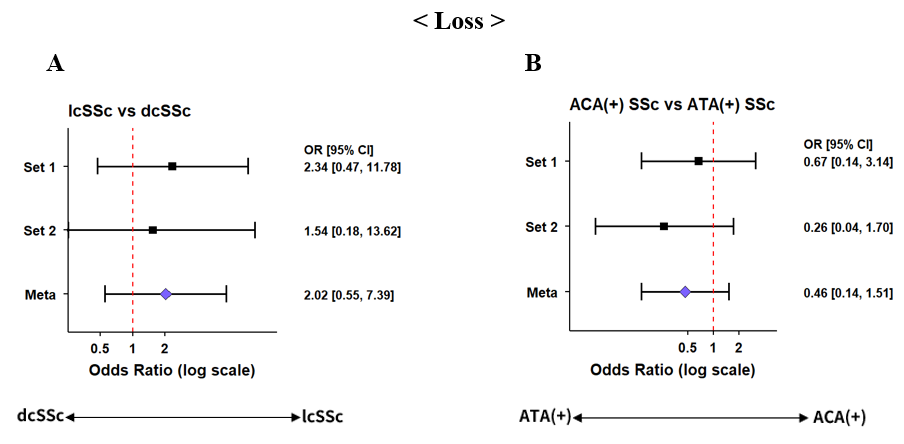

**Supplementary Figure S5. Intra-case associations between Loss and major clinical subtypes of SSc.**

Associations between skin subtypes (lcSSc and dcSSc) and Loss (A) and between autoantibody profiles (ACA and ATA) and Loss (B) are presented. Intra-case results of each dataset and the meta-analysis results are shown. Odds ratios (ORs) are presented on a log scale, and error bars indicate the 95% confidence intervals (CIs). SSc, systemic sclerosis; lcSSc, limited cutaneous SSc; dcSSc, diffuse cutaneous SSc; ACA, anti-centromere antibody; ATA, anti–topoisomerase I antibody.

|  | | **Age category** | **Carriers/Participants** | | | | |
| --- | --- | --- | --- | --- | --- | --- | --- |
|  |  |  | **mCA** | **Loss** | **LOH** | **Gain** | **mLOX** |
| **Set1** | **SSc** | **<50** | **4/241 (1.7%)** | **1/241 (0.4%)** | **3/241 (1.2%)** | **0/241 (0.0%)** | **17/216 (7.9%)** |
|  |  | **50-59** | **3/131 (2.3%)** | **2/131 (1.5%)** | **0/131 (0.0%)** | **1/131 (0.8%)** | **14/116 (12.1%)** |
|  |  | **≥60** | **19/263 (7.2%)** | **8/263 (3.0%)** | **7/263 (2.7%)** | **1/263 (0.4%)** | **45/234 (19.2%)** |
|  | **Control** | **<50** | **19/1286 (1.5%)** | **5/1286 (0.4%)** | **10/1286 (0.8%)** | **4/1286 (0.3%)** | **33/805 (4.1%)** |
|  |  | **50-59** | **58/1323 (4.4%)** | **15/1323 (1.1%)** | **36/1323 (2.7%)** | **3/1323 (0.2%)** | **55/541 (10.2%)** |
|  |  | **≥60** | **112/1792 (6.2%)** | **28/1792 (1.6%)** | **65/1792 (3.6%)** | **12/1792 (0.7%)** | **90/503 (17.9%)** |
| **Set2** | **SSc** | **<50** | **1/62 (1.6%)** | **0/62 (0.0%)** | **0/62 (0.0%)** | **0/62 (0.0%)** | **1/47 (2.1%)** |
|  |  | **50-59** | **1/66 (1.5%)** | **0/66 (0.0%)** | **0/66 (0.0%)** | **0/66 (0.0%)** | **4/58 (6.9%)** |
|  |  | **≥60** | **15/219 (6.8%)** | **6/219 (2.7%)** | **8/219 (3.7%)** | **2/219 (0.9%)** | **23/191 (12.0%)** |
|  | **Control** | **<50** | **7/634 (1.1%)** | **2/634 (0.3%)** | **4/634 (0.6%)** | **3/634 (0.5%)** | **14/400 (3.5%)** |
|  |  | **50-59** | **21/648 (3.2%)** | **11/648 (1.7%)** | **10/648 (1.5%)** | **1/648 (0.2%)** | **22/262 (8.4%)** |
|  |  | **≥60** | **45/888 (5.1%)** | **10/888 (1.1%)** | **27/888 (3.0%)** | **2/888 (0.2%)** | **44/249 (17.7%)** |

**Supplementary Table S1. Age-stratified prevalence of mosaic chromosomal alterations in SSc**

**and controls in two datasets.**

SSc, systemic sclerosis; mCA, mosaic chromosomal alteration; LOH, Loss of heterozygosity; mLOX, mosaic loss of chromosome X.

**Supplementary Table S2. Case–control associations between mosaic chromosomal alterations (mCAs) and SSc in two datasets and meta-analyses.**

| **CaseCtrl** | **Set1** | | | |  | **Set2** | | | |  | **Meta-analysis** | |
| --- | --- | --- | --- | --- | --- | --- | --- | --- | --- | --- | --- | --- |
|  | **Carriers/Participants** | | **OR (95% CI)** | **P** |  | **Carriers/Participants** | | **OR (95% CI)** | **P** |  | **OR (95% CI)** | **P** |
|  | **SSc** | **Control** |  |  |  | **SSc** | **Control** |  |  |  |  |  |
| **mCA** | **26/635 (4.1%)** | **189/4401 (4.3%)** | **1.27 (0.81-2.00)** | **0.30** |  | **17/347 (4.9%)** | **73/2170 (3.4%)** | **1.24 (0.67-2.29)** | **0.50** |  | **1.26 (0.87-1.82)** | **0.22** |
| **Loss** | **11/635 (1.7%)** | **48/4401 (1.1%)** | **2.00 (0.96-4.15)** | **0.065** |  | **6/347 (1.7%)** | **23/2170 (1.1%)** | **1.26 (0.44-3.64)** | **0.67** |  | **1.72 (0.94-3.14)** | **0.079** |
| **LOH** | **10/635 (1.6%)** | **111/4401 (2.5%)** | **0.80 (0.40-1.58)** | **0.52** |  | **8/347 (2.3%)** | **41/2170 (1.9%)** | **0.77 (0.33-1.81)** | **0.55** |  | **0.79 (0.46-1.35)** | **0.38** |
| **Gain** | **2/635 (0.3%)** | **19/4401 (0.4%)** | **1.26 (0.26-6.09)** | **0.77** |  | **2/347 (0.6%)** | **6/2170 (0.3%)** | **1.11 (0.10-12.89)** | **0.93** |  | **1.22 (0.32-4.57)** | **0.77** |
| **mLOX** | **76/566 (13.4%)** | **178/1849 (9.6%)** | **1.31 (0.98-1.75)** | **0.073** |  | **28/296 (9.5%)** | **80/911 (8.8%)** | **0.59 (0.36-0.96)** | **0.034** |  | **1.06 (0.82-1.36)** | **0.65** |

SSc, systemic sclerosis; mCA, mosaic chromosomal alteration; LOH, Loss of heterozygosity; mLOX, mosaic loss of chromosome X; OR, odds ratio; CI, confidence interval.

|  | **Age category** | **Set1** | | | |  | **Set2** | | | |  | **Meta-analysis** | |
| --- | --- | --- | --- | --- | --- | --- | --- | --- | --- | --- | --- | --- | --- |
|  |  | **Carriers/Participants** | | **OR (95% CI)** | **P** |  | **Carriers/Participants** | | **OR (95% CI)** | **P** |  | **OR (95% CI)** | **P** |
|  |  | **SSc** | **Control** |  |  |  | **SSc** | **Control** |  |  |  |  |  |
| **Loss** | **<60** | **3/372 (0.81%)** | **20/2609 (0.77%)** | **1.19 (0.34–4.21)** | **0.79** |  | **0/128 (0.00%)** | **13/1282 (1.01%)** | **0.36 (0.00–2.94)** | **0.42** |  | **1.03 (0.32–3.40)** | **0.96** |
|  | **≥60** | **8/263 (3.04%)** | **28/1792 (1.56%)** | **3.03 (1.11–8.30)** | **0.031** |  | **6/219 (2.74%)** | **10/888 (1.13%)** | **3.00 (0.83–10.76)** | **0.092** |  | **3.02 (1.37–6.66)** | **0.0063** |

**Supplementary Table S3. Age-stratified, case–control associations between Loss and SSc in two datasets and meta-analyses.**

SSc, systemic sclerosis; OR, odds ratio; CI, confidence interval.

**Supplementary Table S4. Case-control associations between Loss and SSc stratified by skin phenotypes in two datasets and meta-analyses.**

| **CaseCtrl** | **Phenotype** | **Set1** | | | |  | **Set2** | | | |  | **Meta-analysis** | |
| --- | --- | --- | --- | --- | --- | --- | --- | --- | --- | --- | --- | --- | --- |
|  |  | **Carriers/Participants** | | **OR (95% CI)** | **P** |  | **Carriers/Participants** | | **OR (95% CI)** | **P** |  | **OR (95% CI)** | **P** |
|  |  | **SSc** | **Control** |  |  |  | **SSc** | **Controls** |  |  |  |  |  |
| **Loss** | **lcSSc** | **9/381 (2.4%)** | **48/4401 (1.1%)** | **2.50 (1.11-5.64)** | **0.028** |  | **5/231 (2.2%)** | **23/2170 (1.1%)** | **1.76 (0.55-5.61)** | **0.34** |  | **2.22 (1.14-4.33)** | **0.019** |
|  | **dcSSc** | **2/254 (0.8%)** |  | **1.08 (0.25-4.61)** | **0.92** |  | **1/115 (0.9%)** |  | **1.06 (0.14-8.18)** | **0.96** |  | **1.07 (0.33-3.50)** | **0.91** |

SSc, systemic sclerosis; lcSSc, limited cutaneous SSc; dcSSc, diffuse cutaneous SSc; OR, odds ratio; CI, confidence interval.

SSc, systemic sclerosis; dcSSc, diffuse cutaneous SSc; lcSSc, limited cutaneous SSc; mLOX, mosaic loss of chromosome X; OR, odds ratio; CI, confidence interval

**Supplementary Table S5. Case-control associations between Loss and SSc stratified by autoantibodies in**

**two datasets and meta-analyses.**

| **CaseCtrl** | **Antibody** | **Set1** | | | |  | **Set2** | | | |  | **Meta-analysis** | |
| --- | --- | --- | --- | --- | --- | --- | --- | --- | --- | --- | --- | --- | --- |
|  |  | **Carriers/Participants** | | **OR (95% CI)** | **P** |  | **Carriers/Participants** | | **OR (95% CI)** | **P** |  | **OR (95% CI)** | **P** |
|  |  | **SSc** | **Control** |  |  |  | **SSc** | **Control** |  |  |  |  |  |
| **Loss** | **ACA(+)** | **6/225 (2.67%)** | **48/4401 (1.09%)** | **2.58 (0.96-6.92)** | **0.06** |  | **3/169 (1.78%)** | **23/2170 (1.06%)** | **1.73 (0.40-7.38)** | **0.46** |  | **2.27 (1.01-5.14)** | **0.049** |
|  | **ATA(+)** | **3/161 (1.86%)** |  | **2.65 (0.78-8.97)** | **0.12** |  | **2/76 (2.63%)** |  | **3.54 (0.77-16.40)** | **0.11** |  | **2.97 (1.14-7.70)** | **0.026** |

SSc, systemic sclerosis; ACA, anti-centromere antibody; ATA, anti–topoisomerase I antibody; OR, odds ratio; CI, confidence interval.

**Supplementary Table S6. Case-control associations between Loss and SSc stratified by clinical manifestations (ILD and VC) in two datasets and meta-analyses.**

| **CaseCtrl** | **Clinical manifestation** | **Set1** | | | |  | **Set2** | | | |  | **Meta-analysis** | |
| --- | --- | --- | --- | --- | --- | --- | --- | --- | --- | --- | --- | --- | --- |
|  |  | **Carriers/Participants** | | **OR (95% CI)** | **P** |  | **Carriers/Participants** | | **OR (95% CI)** | **P** |  | **OR (95% CI)** | **P** |
|  |  | **SSc** | **Control** |  |  |  | **SSc** | **Control** |  |  |  |  |  |
| **Loss** | **ILD(+)** | **6/289 (2.08%)** | **48/4401 (1.09%)** | **2.59 (1.04-6.43)** | **0.040** |  | **3/143 (2.10%)** | **23/2170 (1.06%)** | **2.16 (0.59-7.94)** | **0.24** |  | **2.44 (1.16-5.14)** | **0.019** |
|  | **ILD(-)** | **4/279 (1.43%)** |  | **1.71 (0.57-5.07)** | **0.34** |  | **3/204 (1.47%)** |  | **1.36 (0.35-5.26)** | **0.66** |  | **1.56 (0.67-3.65)** | **0.30** |
|  | **VC(+)** | **6/176 (3.41%)** |  | **4.00 (1.60-10.05)** | **0.0031** |  | **1/88 (1.14%)** |  | **1.22 (0.15-9.85)** | **0.85** |  | **3.30 (1.42-7.66)** | **0.0054** |
|  | **VC(-)** | **4/385 (1.04%)** |  | **1.28 (0.44-3.77)** | **0.65** |  | **5/259 (1.93%)** |  | **1.77 (0.59-5.31)** | **0.31** |  | **1.50 (0.69-3.24)** | **0.30** |

SSc, systemic sclerosis; ILD, interstitial lung disease; VC, vascular complications; OR, odds ratio; CI, confidence interval.

| **CaseCtrl** | | **Conditioned on** | **Set1** | | | |  | **Set2** | | | |  | **Meta-analysis** | |
| --- | --- | --- | --- | --- | --- | --- | --- | --- | --- | --- | --- | --- | --- | --- |
|  |  |  | **Carriers/Participants** | | **OR (95% CI)** | **P** |  | **Carriers/Participants** | | **OR (95% CI)** | **P** |  | **OR (95% CI)** | **P** |
|  |  |  | **SSc** | **Control** |  |  |  | **SSc** | **Control** |  |  |  |  |  |
| **Loss** | **ILD(+)** | **ACA(Set1:40, Set2:23)** | **6/289 (2.08%)** | **48/4401 (1.09%)** | **2.94 (1.03-7.01)** | **0.045** |  | **3/143 (2.10%)** | **23/2170 (1.06%)** | **2.95 (0.74-8.98)** | **0.11** |  | **2.95 (1.38-6.31)** | **0.0054** |
|  |  | **ATA(Set1:13, Set2:6)** |  |  | **2.70 (0.71-7.46)** | **0.13** |  |  |  | **2.72 (0.22-9.03)** | **0.44** |  | **2.52 (0.93-6.79)** | **0.068** |
|  |  | **lcSSc(Set1:98, Set2:62)** |  |  | **1.72 (0.34-5.36)** | **0.45** |  |  |  | **2.19 (0.22-9.26)** | **0.42** |  | **1.88 (0.62-5.68)** | **0.27** |
|  | **VC(+)** | **ACA(Set1:52, Set2:32)** | **6/176 (3.41%)** |  | **2.75 (0.54-8.61)** | **0.19** |  | **1/88 (1.14%)** |  | **0.99 (0.00-7.81)** | **0.99** |  | **2.58 (0.68-9.87)** | **0.17** |
|  |  | **ATA(Set1:69, Set2:26)** |  |  | **4.49 (1.36-11.92)** | **0.017** |  |  |  | **2.27 (0.22-10.90)** | **0.43** |  | **3.81 (1.48-9.85)** | **0.0057** |
|  |  | **lcSSc(Set1:81, Set2:45)** |  |  | **2.00 (0.18-7.85)** | **0.47** |  |  |  | **1.29 (0.00-10.11)** | **0.87** |  | **1.92 (0.32-11.53)** | **0.48** |

SSc, systemic sclerosis; lcSSc, limited cutaneous SSc; ACA, anti-centromere antibody; ATA, anti–topoisomerase I antibody; ILD, interstitial lung disease; VC, vascular complications; OR, odds ratio; CI, confidence interval.

**Supplementary Table S7. Case–control associations between Loss and ILD(+) SSc or VC(+) SSc conditioned on antibody profiles or skin phenotype, in two datasets and meta-analyses.**

| **CaseCtrl** | | | **Set1** | | | |  | **Set2** | | | |  | **Meta-analysis** | |
| --- | --- | --- | --- | --- | --- | --- | --- | --- | --- | --- | --- | --- | --- | --- |
|  |  |  | **Carriers/Participants** | | **OR (95% CI)** | **P** |  | **Carriers/Participants** | | **OR (95% CI)** | **P** |  | **OR (95% CI)** | **P** |
|  |  |  | **SSc** | **Control** |  |  |  | **SSc** | **Control** |  |  |  |  |  |
| **CF> 5%** | **Loss** | **SSc** | **8/632 (1.3%)** | **27/4380 (0.6%)** | **2.63 (1.07–6.47)** | **0.035** |  | **6/347 (1.7%)** | **19/2166 (0.9%)** | **1.70 (0.59–4.93)** | **0.33** |  | **2.20 (1.11–4.36)** | **0.025** |
|  |  | **lcSSc** | **6/378 (1.6%)** |  | **2.89 (1.04–8.02)** | **0.042** |  | **5/231 (2.2%)** |  | **1.99 (0.60–6.57)** | **0.26** |  | **2.47 (1.14–5.37)** | **0.023** |
|  |  | **ACA(+)** | **5/224 (2.2%)** |  | **3.90 (1.25–12.21)** | **0.019** |  | **3/169 (1.8%)** |  | **1.94 (0.44–8.67)** | **0.39** |  | **3.02 (1.22–7.48)** | **0.017** |
|  |  | **ILD(+)** | **4/287 (1.4%)** |  | **3.21 (1.04–9.96)** | **0.043** |  | **3/143 (2.1%)** |  | **2.52 (0.67–9.48)** | **0.17** |  | **2.90 (1.23–6.86)** | **0.015** |
|  |  | **VC(+)** | **5/175 (2.9%)** |  | **6.15 (2.15–17.59)** | **0.00070** |  | **1/88 (1.1%)** |  | **1.41 (0.17–11.64)** | **0.75** |  | **4.59 (1.79–11.77)** | **0.0015** |

SSc, systemic sclerosis; lcSSc, limited cutaneous SSc; ACA, anti-centromere antibody; ILD, interstitial lung disease; VC, vascular complications; OR, odds ratio; CI, confidence interval; CF, cell fraction.

**Supplementary Table S8. Case–control associations between Loss with high–cell-fraction (CF> 5%) and SSc subtypes in two datasets and meta-analyses.**

| **CaseCtrl** | | | **Set1** | | | |  | **Set2** | | | |  | **Meta-analysis** | |
| --- | --- | --- | --- | --- | --- | --- | --- | --- | --- | --- | --- | --- | --- | --- |
|  |  |  | **Carriers/Participants** | | **OR (95% CI)** | **P** |  | **Carriers/Participants** | | **OR (95% CI)** | **P** |  | **OR (95% CI)** | **P** |
|  |  |  | **SSc** | **Control** |  |  |  | **SSc** | **Control** |  |  |  |  |  |
| **No CF threshold** | **mLOX** | **SSc** | **76/566 (13.4%)** | **178/1849 (9.6%)** | **1.31 (0.98-1.75)** | **0.073** |  | **28/296 (9.5%)** | **80/911 (8.8%)** | **0.59 (0.36-0.96)** | **0.034** |  | **1.06 (0.82-1.36)** | **0.65** |
|  |  | **lcSSc** | **61/353 (17.3%)** |  | **1.57 (1.13-2.17)** | **0.007** |  | **28/203 (13.8%)** |  | **0.80 (0.48-1.33)** | **0.39** |  | **1.29 (0.98-1.70)** | **0.07** |
|  |  | **ACA(+)** | **44/214 (20.56%)** |  | **1.62 (1.10-2.37)** | **0.014** |  | **27/156 (17.31%)** |  | **1.04 (0.61-1.77)** | **0.88** |  | **1.39 (1.02-1.90)** | **0.037** |
| **CF> 5%** | **mLOX** | **SSc** | **10/500 (2.00%)** | **12/1683 (0.71%)** | **2.39 (1.02-5.60)** | **0.045** |  | **2/270 (0.74%)** | **3/834 (0.36%)** | **1.38 (0.22-8.68)** | **0.73** |  | **2.17 (1.00-4.70)** | **0.05** |
|  |  | **lcSSc** | **9/301 (2.99%)** |  | **3.04 (1.25-7.40)** | **0.014** |  | **2/177 (1.13%)** |  | **2.09 (0.32-13.59)** | **0.44** |  | **2.84 (1.27-6.34)** | **0.011** |
|  |  | **ACA(+)** | **8/178 (4.49%)** |  | **3.96 (1.54-10.19)** | **0.0043** |  | **2/131 (1.53%)** |  | **2.90 (0.44-19.04)** | **0.27** |  | **3.72 (1.60-8.65)** | **0.0023** |

SSc, systemic sclerosis; lcSSc, limited cutaneous SSc; ACA, anti-centromere antibody; mLOX, mosaic loss of chromosome X; OR, odds ratio; CI, confidence interval; CF, cell fraction.

**Supplementary Table S10. Intra-case associations between mLOX and SSc stratified by**

**skin phenotypes or autoantibodies in two datasets and meta-analyses.**

| **IntraCase** | **Set1** | | | |  | **Set2** | | | |  | **Meta-analysis** | |
| --- | --- | --- | --- | --- | --- | --- | --- | --- | --- | --- | --- | --- |
|  | **Carriers/Participants** | | **OR (95% CI)** | **P** |  | **Carriers/Participants** | | **OR (95% CI)** | **P** |  | **OR (95% CI)** | **P** |
|  | **lcSSc** | **dcSSc** |  |  |  | **lcSSc** | **dcSSc** |  |  |  |  |  |
| **mLOX** | **61/353 (17.28%)** | **15/213 (7.04%)** | **2.32 (1.27-4.27)** | **0.0065** |  | **28/203 (13.79%)** | **0/92 (0.00%)** | **23.55 (3.20-3004.20)** | **0.00015** |  | **2.49 (1.37-4.54)** | **0.0027** |

| **IntraCase** | **Set1** | | | |  | **Set2** | | | |  | **Meta-analysis** | |
| --- | --- | --- | --- | --- | --- | --- | --- | --- | --- | --- | --- | --- |
|  | **Carriers/Participants** | | **OR (95% CI)** | **P** |  | **Carriers/Participants** | | **OR (95% CI)** | **P** |  | **OR (95% CI)** | **P** |
|  | **ACA(+)** | **ATA(+)** |  |  |  | **ACA(+)** | **ATA(+)** |  |  |  |  |  |
| **mLOX** | **43/206 (20.87%)** | **9/128 (7.03%)** | **2.74 (1.23-6.13)** | **0.014** |  | **26/151 (17.22%)** | **1/59 (1.69%)** | **8.03 (1.02-63.48)** | **0.048** |  | **3.16 (1.49-6.68)** | **0.0026** |

SSc, systemic sclerosis; lcSSc, limited cutaneous SSc; dcSSc, diffuse cutaneous SSc; ACA, anti-centromere antibody; ATA, anti–topoisomerase I antibody; mLOX, mosaic loss of chromosome X; OR, odds ratio; CI, confidence interval.

| **IntraCase** | | **Set1** | | | |  | **Set2** | | | |  | **Meta-analysis** | |
| --- | --- | --- | --- | --- | --- | --- | --- | --- | --- | --- | --- | --- | --- |
|  |  | **Carriers/Participants** | | **OR (95% CI)** | **P** |  | **Carriers/Participants** | | **OR (95% CI)** | **P** |  | **OR (95% CI)** | **P** |
|  |  | **lcSSc** | **dcSSc** |  |  |  | **lcSSc** | **dcSSc** |  |  |  |  |  |
| **CF> 5%** | **mLOX** | **9/301 (2.99%)** | **1/199 (0.50%)** | **3.79 (0.47-30.51)** | **0.21** |  | **2/177 (1.13%)** | **0/92 (0.00%)** | **2.75 (0.22-381.07)** | **0.47** |  | **3.51 (0.57-21.68)** | **0.18** |

**Supplementary Table S11. Intra–case associations between high–cell-fraction (CF> 5%) mLOX and SSc subtypes in two datasets and meta-analyses.**

| **IntraCase** | | **Set1** | | | |  | **Set2** | | | |  | **Meta-analysis** | |
| --- | --- | --- | --- | --- | --- | --- | --- | --- | --- | --- | --- | --- | --- |
|  |  | **Carriers/Participants** | | **OR (95% CI)** | **P** |  | **Carriers/Participants** | | **OR (95% CI)** | **P** |  | **OR (95% CI)** | **P** |
|  |  | **ACA(+)** | **ATA(+)** |  |  |  | **ACA(+)** | **ATA(+)** |  |  |  |  |  |
| **CF> 5%** | **mLOX** | **8/178 (4.49%)** | **0/126 (0.00%)** | **6.95 (0.82-907.50)** | **0.082** |  | **2/131 (1.53%)** | **0/62 (0.00%)** | **2.50 (0.20-347.77)** | **0.52** |  | **4.31 (0.34-55.43)** | **0.26** |

SSc, systemic sclerosis; lcSSc, limited cutaneous SSc; dcSSc, diffuse cutaneous SSc; ACA, anti-centromere antibody; ATA, anti–topoisomerase I antibody; mLOX, mosaic loss of chromosome X; OR, odds ratio; CI, confidence interval; CF, cell fraction.

| **IntraCase** | **Set1** | | | |  | **Set2** | | | |  | **Meta-analysis** | |
| --- | --- | --- | --- | --- | --- | --- | --- | --- | --- | --- | --- | --- |
|  | **Carriers/Participants** | | **OR (95% CI)** | **P** |  | **Carriers/Participants** | | **OR (95% CI)** | **P** |  | **OR (95% CI)** | **P** |
|  | **lcSSc** | **dcSSc** |  |  |  | **lcSSc** | **dcSSc** |  |  |  |  |  |
| **Loss** | **9/381 (2.36%)** | **2/254 (0.79%)** | **2.34 (0.47-11.78)** | **0.30** |  | **5/231 (2.16%)** | **1/115 (0.87%)** | **1.54 (0.18-13.62)** | **0.70** |  | **2.02 (0.55-7.39)** | **0.29** |

| **IntraCase** | **Set1** | | | |  | **Set2** | | | |  | **Meta-analysis** | |
| --- | --- | --- | --- | --- | --- | --- | --- | --- | --- | --- | --- | --- |
|  | **Carriers/Participants** | | **OR (95% CI)** | **P** |  | **Carriers/Participants** | | **OR (95% CI)** | **P** |  | **OR (95% CI)** | **P** |
|  | **ACA(+)** | **ATA(+)** |  |  |  | **ACA(+)** | **ATA(+)** |  |  |  |  |  |
| **Loss** | **6/225 (2.67%)** | **3/161 (1.86%)** | **0.67 (0.14-3.14)** | **0.61** |  | **3/169 (1.78%)** | **2/76 (2.63%)** | **0.26 (0.040-1.70)** | **0.16** |  | **0.46 (0.14-1.51)** | **0.20** |

SSc, systemic sclerosis; lcSSc, limited cutaneous SSc; dcSSc, diffuse cutaneous SSc; ACA, anti-centromere antibody; ATA, anti–topoisomerase I antibody; OR, odds ratio; CI, confidence interval.

**Supplementary Table S12. Intra-case associations between Loss and SSc stratified by**

**skin phenotypes or autoantibodies in two datasets and meta-analyses.**

**Supplementary Table S13. Case–control associations between Loss and late-onset SSc subsets in two datasets and meta-analyses.**

| **CaseCtrl** | **Subsets** | **Set1** | | | |  | **Set2** | | | |  | **Meta-analysis** | |
| --- | --- | --- | --- | --- | --- | --- | --- | --- | --- | --- | --- | --- | --- |
|  |  | **Carriers/Participants** | | **OR (95% CI)** | **P** |  | **Carriers/Participants** | | **OR (95% CI)** | **P** |  | **OR (95% CI)** | **P** |
|  |  | **Late-onset SSc** | **Control** |  |  |  | **Late-onset SSc** | **Control** |  |  |  |  |  |
| **Loss** | **Overall** | **4/56 (7.1%)** | **48/4401 (1.1%)** | **5.85 (1.64-20.85)** | **0.0064** |  | **2/121 (1.7%)** | **23/2170 (1.1%)** | **1.97 (0.37-10.58)** | **0.43** |  | **3.94 (1.43-10.86)** | **0.0080** |
|  | **lcSSc** | **3/40 (7.5%)** |  | **6.97 (1.43-34.07)** | **0.017** |  | **2/89 (2.2%)** |  | **3.05 (0.54-17.15)** | **0.21** |  | **4.77 (1.48-15.35)** | **0.0088** |
|  | **ACA(+)** | **3/24 (12.5%)** |  | **17.73 (2.95-106.71)** | **0.0017** |  | **1/66 (1.5%)** |  | **2.11 (0.20-22.40)** | **0.54** |  | **8.13 (1.95-33.96)** | **0.0040** |
|  | **ILD(+)** | **1/28 (3.6%)** |  | **4.10 (0.51-32.82)** | **0.18** |  | **1/47 (2.1%)** |  | **3.06 (0.36-26.25)** | **0.31** |  | **3.56 (0.80-15.86)** | **0.096** |
|  | **VC(+)** | **2/16 (12.5%)** |  | **7.99 (1.39-45.93)** | **0.020** |  | **0/26 (0.0%)** |  | **2.70 (0.02-29.04)** | **0.57** |  | **6.53 (1.35-31.60)** | **0.020** |

SSc, systemic sclerosis; lcSSc, limited cutaneous SSc; ACA, anti-centromere antibody; ILD, interstitial lung disease ; VC, vascular complications; OR, odds ratio; CI, confidence interval.

**Supplementary Table S14. Case–control associations between Loss and Non-late-onset SSc subsets in two datasets and meta-analyses.**

| **CaseCtrl** | **Subsets** | **Set1** | | | |  | **Set2** | | | |  | **Meta-analysis** | |
| --- | --- | --- | --- | --- | --- | --- | --- | --- | --- | --- | --- | --- | --- |
|  |  | **Carriers/Participants** | | **OR (95% CI)** | **P** |  | **Carriers/Participants** | | **OR (95% CI)** | **P** |  | **OR (95% CI)** | **P** |
|  |  | **Non-late-onset SSc** | **Control** |  |  |  | **Non-late-onset SSc** | **Control** |  |  |  |  |  |
| **Loss** | **Overall** | **1/132 (0.8%)** | **48/4401 (1.1%)** | **0.96 (0.13-7.20)** | **0.97** |  | **4/222 (1.8%)** | **23/2170 (1.1%)** | **1.97 (0.62-6.33)** | **0.25** |  | **1.65 (0.60-4.52)** | **0.33** |
|  | **lcSSc** | **1/100 (1.0%)** |  | **1.25 (0.17-9.46)** | **0.83** |  | **3/138 (2.2%)** |  | **2.18 (0.57-8.39)** | **0.26** |  | **1.84 (0.60-5.64)** | **0.29** |
|  | **ACA(+)** | **1/45 (2.2%)** |  | **2.69 (0.34-21.02)** | **0.35** |  | **2/100 (2.0%)** |  | **2.33 (0.46-11.92)** | **0.31** |  | **2.46 (0.69-8.84)** | **0.17** |
|  | **ILD(+)** | **0/48 (0.0%)** |  | **1.34 (0.01-10.00)** | **0.85** |  | **2/95 (2.1%)** |  | **2.62 (0.57-12.09)** | **0.22** |  | **2.35 (0.58-9.47)** | **0.23** |
|  | **VC(+)** | **1/32 (3.1%)** |  | **4.71 (0.61-36.63)** | **0.14** |  | **1/62 (1.6%)** |  | **1.87 (0.24-14.85)** | **0.55** |  | **2.98 (0.69-12.80)** | **0.14** |

SSc, systemic sclerosis; lcSSc, limited cutaneous SSc; ACA, anti-centromere antibody; ILD, interstitial lung disease ; VC, vascular complications; OR, odds ratio; CI, confidence interval.

| **CaseCtrl** | **Subsets** | **Set1** | | | |  | **Set2** | | | |  | **Meta-analysis** | |
| --- | --- | --- | --- | --- | --- | --- | --- | --- | --- | --- | --- | --- | --- |
|  |  | **Carriers/Participants** | | **OR (95% CI)** | **P** |  | **Carriers/Participants** | | **OR (95% CI)** | **P** |  | **OR (95% CI)** | **P** |
|  |  | **Late-onset SSc** | **Control** |  |  |  | **Late-onset SSc** | **Control** |  |  |  |  |  |
| **mLOX** | **Overall** | **14/46 (30.4%)** | **178/1849 (9.6%)** | **2.00 (1.01–3.98)** | **0.047** |  | **12/102 (11.8%)** | **80/911 (8.8%)** | **0.33 (0.15–0.76)** | **0.0089** |  | **0.96 (0.57–1.63)** | **0.88** |
|  | **lcSSc** | **12/36 (33.3%)** |  | **2.32 (1.09–4.94)** | **0.029** |  | **12/77 (15.6%)** |  | **0.47 (0.20–1.08)** | **0.074** |  | **1.12 (0.64–1.97)** | **0.68** |
|  | **ACA(+)** | **9/23 (39.1%)** |  | **3.05 (1.23–7.54)** | **0.016** |  | **11/59 (18.6%)** |  | **0.61 (0.25–1.45)** | **0.26** |  | **1.31 (0.70–2.46)** | **0.39** |

SSc, systemic sclerosis; lcSSc, limited cutaneous SSc; ACA, anti-centromere antibody; mLOX, mosaic loss of chromosome X; OR, odds ratio; CI, confidence interval.

**Supplementary Table S15. Case–control associations between mLOX and late-onset SSc subsets in two datasets and meta-analyses.**

**Supplementary Table S16. Case–control associations between mLOX and Non-late-onset SSc subsets in two datasets and meta-analyses.**

|  |  | **Set1** | | | |  | **Set2** | | | |  | **Meta-analysis** | |
| --- | --- | --- | --- | --- | --- | --- | --- | --- | --- | --- | --- | --- | --- |
| **CaseCtrl** | **Subsets** | **Carriers/Participants** | | **OR (95% CI)** | **P** |  | **Carriers/Participants** | | **OR (95% CI)** | **P** |  | **OR (95% CI)** | **P** |
|  |  | **Non-late-onset SSc** | **Control** |  |  |  | **Non-late-onset SSc** | **Control** |  |  |  |  |  |
| **mLOX** | **SSc** | **19/117 (16.2%)** | **178/1849 (9.6%)** | **1.72 (1.01–2.92)** | **0.045** |  | **16/190 (8.4%)** | **80/911 (8.8%)** | **0.68 (0.38–1.21)** | **0.19** |  | **1.12 (0.76–1.66)** | **0.56** |
|  | **lcSSc** | **17/92 (18.5%)** |  | **1.91 (1.08–3.36)** | **0.026** |  | **16/122 (13.1%)** |  | **0.98 (0.54–1.78)** | **0.94** |  | **1.39 (0.92–2.10)** | **0.12** |
|  | **ACA** | **12/43 (27.9%)** |  | **2.66 (1.31–5.39)** | **0.0068** |  | **16/94 (17.0%)** |  | **1.29 (0.70–2.38)** | **0.42** |  | **1.76 (1.10–2.79)** | **0.017** |

SSc, systemic sclerosis; lcSSc, limited cutaneous SSc; ACA, anti-centromere antibody; mLOX, mosaic loss of chromosome X; OR, odds ratio; CI, confidence interval.
